# Rigorous software pipeline for clinical somatic mutation analyses of solid tumors

**DOI:** 10.1101/2023.06.08.23291143

**Authors:** Ivaylo Stoimenov, Marina Rashyna, Tom Adlerteg, Luís Nunes, Joakim Ekström, Viktor Ljungström, Lucy Mathot, Ian Cheong, Tobias Sjöblom

## Abstract

Mutational analyses of tumor DNA guide the use of targeted therapies and checkpoint inhibitors in management of solid tumors. Reducing false positive mutation calls without compromising sensitivity as gene panels increase in size, and whole exome and genome sequencing enters clinical use, remains a major challenge. Aiming for robust somatic mutation analyses in the clinical setting, we have developed VARify, an integrated, accurate and computationally efficient software for cancer genome analyses encompassing all steps from pre-processing of sequencing reads to mutation identification. Benchmarking to two state-of-the-art open-source somatic mutation analysis pipelines demonstrated accurate detection of clinically actionable point mutations, all while strongly reducing the number of false positive mutations reported, at comparable or faster speed. Further, the VARify output classified microsatellite unstable colorectal cancers by tumor mutation burden better than the other pipelines. In comparisons where the same tumors were subjected to different panel enrichment and sequencing technologies, VARify had the most consistent intersection of consensus mutations. False positive calls were produced when the same data was used as tumor and reference by the other pipelines, while VARify did not produce such calls. The calling uniformity across sequencing technologies of VARify and its tumor-only analysis derivative pipeline ALTOmate was also demonstrated. Taken together, these two novel pipelines can improve clinical mutation analysis to the benefit of cancer patients.

## Background

The discovery of the genetic basis of cancer spurred the development of oncogene-targeted therapies, with notable examples in sotorasib (KRAS), vemurafenib (BRAF), dacomitinib (EGFR) or ensartinib (ALK) for lung cancer (1) or cetuximab (EGFR) for colorectal cancer (2). Further, checkpoint blockade immunotherapies have entered routine oncology (3, 4). The clinical use of these therapies requires companion diagnostics that can detect somatic mutations with high sensitivity and specificity in patient tumors (4). Typically, tumor tissue used in diagnostic gene sequencing is composed not only of tumor cells but also of other cell types, including normal epithelial, stromal and inflammatory cells, and can therefore have a low tumor cell fraction (5, 6). Samples often fail to meet inclusion criteria based on low tumor cell content and DNA yield, but are still analyzed due to lack of other samples of sufficient quality (7, 8). The combination of insufficient tumor purity, high tumor heterogeneity, and the allelic imbalances in cancer genomes, can result in sequencing data that is difficult to interpret and report mutations from with a high degree of confidence (9, 10). Thus, mutational analyses of tumor tissues have challenges beyond those encountered in analyses of constitutional DNA (11–13).

A systematic review of next-generation sequencing (NGS) in the molecular pathology setting revealed that 83% of successfully analyzed samples had at least one mutation detected by gene panel sequencing of 50 to 500 genes (14). The use of comprehensive gene panels over single gene analysis is cost-effective (15), as improved detection of actionable biomarkers and better therapeutic guidance benefits patients (16). Therefore, targeted gene panels for cancer diagnostics are continuously increasing in size, yielding large amounts of sequencing data covering many genes and detection of many putative mutations of unknown significance. These challenges compound when analyzing whole exomes and whole genomes of tumors. The value of false positive reduction from sequencing also a patient-matched normal DNA sample is supported by recent studies (17, 18), but the vast majority of molecular pathology analyses are still performed using only a tumor sample. In all, clinical sequencing of solid tumors beyond hotspot mutation detection has demands that are partially unmet by the current state-of-art in molecular pathology.

Enrichment technologies coupled with NGS have increased the sensitivity of mutation detection by enabling sampling of genomic regions of interest to thousand-fold depths. The sequenced reads are then trimmed of non-target sequences, for example using Cutadapt (19), and then aligned to a reference genome sequence using software such as BWA (20). Next, one or more of several tools for single nucleotide variant and indel detection, including Strelka2 (21), Mutect2 (22), VarScan2 (23) and others (24), is used to identify variant bases. The output from secondary sequence analysis is then further interpreted by annotation software and molecular diagnostic experts. As clinical sequencing now matures into a routine diagnostic activity, increasing regulatory authority demands create a need for validated software pipelines that integrate the tasks of read trimming, alignment, and mutation calling (25). These functionalities are today often performed by independent software packages of different origins combined into a pipeline (26). To ensure that these functionalities perform optimally as a unit, and in compliance with regulatory requirements, there is a need for full integration. It is also important to eliminate unnecessary or untestable assumptions from analysis models to ensure diagnostic performance under any condition and facilitate regulatory approval.

In light of the above, the ideal analysis software should operate from targeted panels to whole exomes and whole genomes, handle sequence data of several different vendor origins including long-read sequencing, show consistency when re-analyzing a sample, and meet requirements for regulatory approval. Here, we describe an integrated solution for read trimming, read alignment and mutation calling developed specifically for clinical sequencing of solid tumors. We benchmark each key functionality and the integrated solution to widely used reference software, and compare mutation detection performance in presence and absence of normal tissue reference.

## Methods

### Tumor and patient-matched normal sequence datasets

The mutational analyses of patient samples were approved by the Ethical Review Board (EPN Uppsala, C116/2007). Genomic DNA from 107 CRC patients (matched tumor and normal specimens) was subjected to target enrichment of the coding sequence of 676 cancer-related genes and gene families using a custom-designed Haloplex panel (Agilent) and sequenced on a HiSeq 2000 platform (Illumina) to >1000-fold average read depth in regions of interest for both tumor and normal samples (27). In addition, DNA from 4 of the 107 T/N pairs was subjected to library preparation using TruSight Oncology 500 and sequenced on NextSeq 550 platform (Illumina), and prepared with Core Exome (Twist Biosciences) library panel followed by sequencing on NovaSeq 6000 (Illumina). The 4 tumor samples were comprised of one microsatellite instability-high (MSI-High), one microsatellite instability-low (MSI-Low) and two microsatellite stable (MSS) samples, to represent all three types of instability phenotypes from the complete benchmarking dataset. The same samples used for comparing different Illumina sequencing platforms were re-sequenced using ion- semiconductor sequencing technology (IonTorrent, Thermo Fisher Scientific). Unfortunately, the paired normal of one case (MSI-High) was no longer available, which limited some parts of the analysis for this sample. All data produced in the present study are available upon reasonable request to the authors.

### Sequencing and Quality Control Phase 2 dataset

The Somatic Mutation Working Group of the SEQC2 consortium, led by the US Food and Drug Administration (FDA), has released datasets for benchmarking somatic mutation calling pipelines (28). The genomic reference sample SRP292966 provided by SEQC2 was accessed from the SRA archive (29). The dataset included several technical repeats of whole-exome sequenced (WES) reference material by three different sequencing protocols based on library preparation kits from Roche Sequencing Solutions, Integrated DNA technologies or Agilent Technologies and subjected to sequencing by ligation (Illumina) (30). Further, there are 220 true positive mutation calls in the SEQC2 dataset, which have all been orthogonally validated by ddPCR (30). From 284 randomly selected putative calls, only 220 are true positives, while the remaining 64 are false positives with the mutation present in the normal tissue, thus constituting germline variants as opposed to somatic mutations (30).

### State-of-the-art software benchmarks

Two commonly used open-sourced pipelines for somatic mutation detection were used for benchmarking. The adapter trimming in both was performed by Cutadapt v.4.0 (19). The trimmed sequencing reads were aligned with BWA-MEM v.0.7.17 over the GRCh37 reference human assembly and the output sam-files were processed with Samtools v.1.15 prior calling. The software used for somatic mutation identification was either Mutect v.2 from GATK 4.1.3 (in the CBSM pipeline, Figure 1A) or Strelka v.2.9.10 (in the CBSS pipeline, Figure 1B). Both pipelines were executed with default settings in somatic mode according to best practices and recommendations in their documentation.

**Figure 1.**
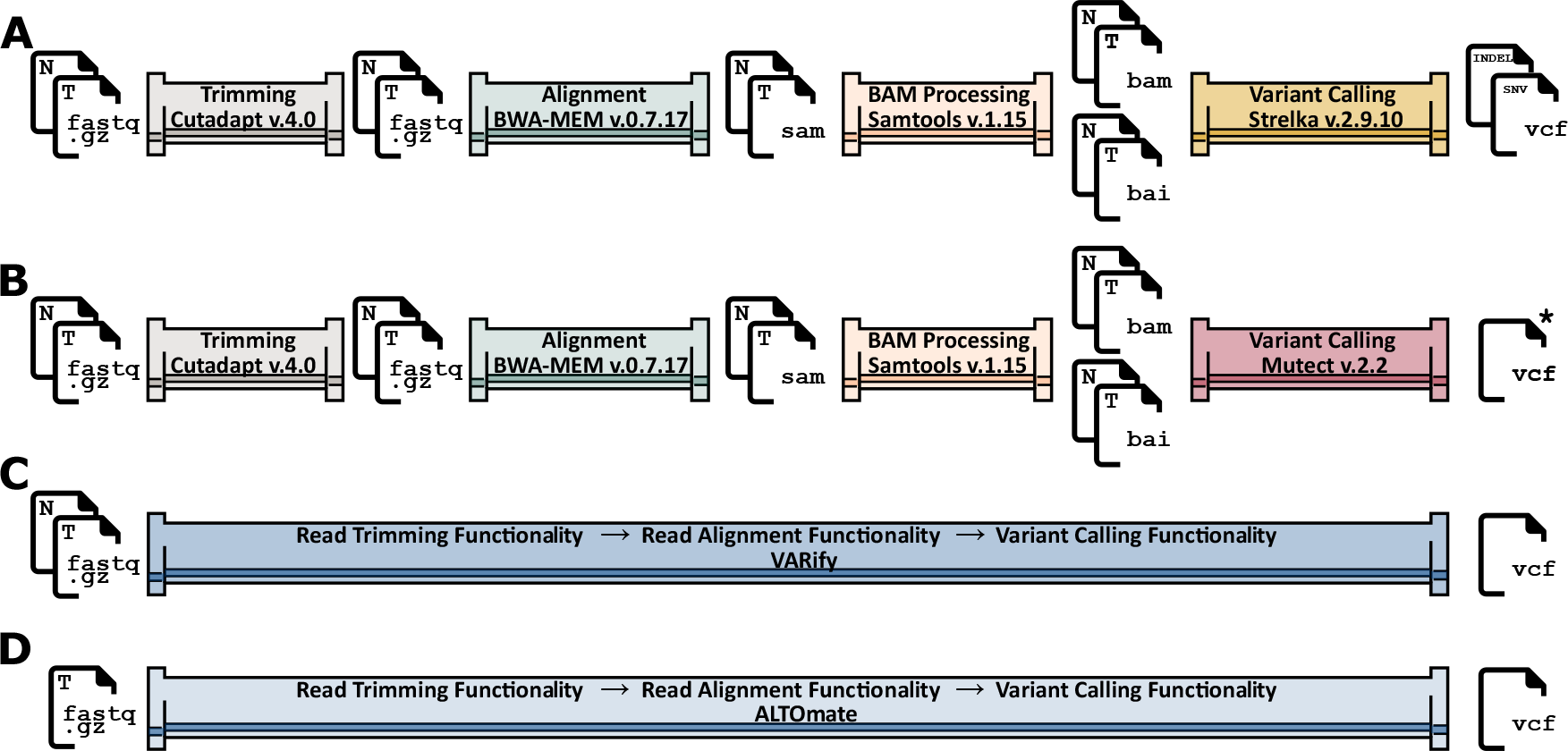
VARify and ALTOmate are integrated high-performance pipelines for mutational analysis in cancer. The four pipelines accept sequencing data input in FASTQ file format and output raw somatic calls in the VCF file format. (**A**) Pipeline based on Cutadapt, BWA-MEM, Samtools and Strelka2 (CBSS) (**B**) Pipeline based on Cutadapt, BWA-MEM, Samtools and Mutect2 (CBSM). (**C**) The VARify pipeline, processing the raw sequencing data of tumor and patient-matched normal sample into a standard variation report. (**D**) The ALTOmate pipeline, processing tumor-only sequencing data into a standard variation report. Intermediate input/output files, generated to interface the separate software applications in CBSS and CBSM are shown. * An additional step of Picard Tools is necessary to filter Mutect2 mutations. T, tumor sample, N, normal sample.

### Novel mutation analysis pipeline

To maximize efficiency, we developed a tightly integrated software incorporating the necessary components of a somatic mutation analysis pipeline in a single package termed VARify. Thus, conceptually discrete steps in the analysis such as the read trimmer and/or cutter, read aligner, variant caller and the necessary input/output operations can be perceived as functionalities, rather than as separate executables. To enhance performance, we minimized intermediate file output/sorting and implemented task parallelism in all the different functionalities.

The leading sequencing platforms (Illumina, MGI, PacBio and IonTorrent) output sequencing data in the *de facto* standard FASTQ file format (31), therefore, the input functionality (IF) supports FASTQ format in single-read or pair-end mode. The read trimming functionality (RTF), provided with an input adapter sequence, performs exact or partial matching of the adapter to the read sequence. If an exact match is not found, a full-length Smith-Waterman gapped alignment of the adapter and the read sequence is performed in an affine gap model. If the gap-tolerant match is supported by >90% identity, a 3′-adapter trimming is performed on the read with minimum trimming length of three nucleotides. The RTF can also be instructed to cut a fixed-length sequence from the 5′- and/or 3′-end of the sequencing read in both single-read and paired-end modes in a read cutting functionality. The resulting sequence post trimming and/or cutting is directly processed in memory by the read-aligner functionality (RAF). The custom hash-based aligner performs indel-tolerant read mapping to the entire human reference genome usually resulting in several potential genome loci for the read placement. For all putative loci, a full-length Smith-Waterman alignment with affine gaps is performed and the top-scoring alignment for the read location is reported. The pipeline is capable of pair-end read alignment or single read mode. To eliminate erroneous support for somatic mutations coming from uncertainties in structural variation of the human reference sequence, the RAF was designed to not report alignment if members of a read pair map to different chromosomes. To achieve full freedom of mapping, the reads in a pair are mapped independently of each other but paired alignments are given priority by the RAF. If one of the reads in a pair fails to map, alignment in the vicinity of the other member of the pair is not enforced, but the aligned read is reported as a singleton. The insertions and deletions events reported by RAF are left aligned to fulfill the criteria for normalization when the variant is reported as fully left-aligned and parsimonious (32). Next, the variant calling functionality (VCF) groups the reads with overlapping coordinates in structures termed read chunks. Each chunk is delimited from the neighboring chunks by genome positions, where there is no coverage of the reference sequence. The mutation calling is performed in chunks by evaluating the probabilities of the base being correctly sequenced and mapped. The base quality score reported by the sequencing instrument is a measure of the probability of the base being correctly sequenced. The mapping quality score of a sequence read is assigned by the RAF to represent the probability of the read to be correctly placed over the genome. The pattern recognition distance metric Poisson-Binomial radius (PBR) is used to perform statistical evaluation of every putative mutation (33). Based on mapping qualities of included reads, the PBR is computed to quantify the non-overlap of the reference allele probability distributions, along with a sequence error corrected approximation of the mean reference allele ratio difference between tumor and normal sample. Given non-identically distributed sets of reference allele probabilities in the matched tumor and normal sample, the PBR value is the number of joint confidence intervals that do not include 0. Thus, a higher PBR value translates to a higher likelihood of a true somatic mutation. The advantage of PBR to calculations of exact confidence intervals of reference allele ratios is computational speed. The difference between the arithmetic means of the reference allele probabilities in tumor and normal sample (ΔM) can be thought of as a sequence error corrected variant allele frequency (VAF) value. VARify by default allows ≤1% mutant allele in the normal sample to avoid discarding true positive mutations because of sequencing errors in the normal sample. The final result from mutational analysis is reported by the output functionality in the standard Variant Calling file format (.vcf) (34).

### Paired-sample test based on identical samples

The sequencing reads of a normal NCI SEQC2 sample (29) (Normal) were copied and the read IDs altered in one symbol to avoid any possible computational mix-up of samples. The sequencing data and base qualities were identical in both samples. The FASTQ copy was labelled altered normal (aNormal) and the pair aNormal-Normal was analyzed as a tumor-normal pair with VARify, CBSM and CBSS pipelines.

### Statistical analyses and data visualization

The categorization of CRC samples into MSS and MSI-High cases, based on the number of mutations was evaluated by Mann-Whitney U test with Bonferroni correction (pandas 1.5.3, matplotlib 3.5.1, seaborn 0.11.2) and the respective p-values were used. Cohen’s d was used to estimate the effect size in each comparison. The ROC analysis of the same experiment was performed in R v.4.1.3 using pROC package v.1.18.0. The Matplotlibv.3.5.1 library in Python v.3.8.10 was used to generate plots. Figures were prepared with Inkscape v.1.2.2.

## Results

Colorectal cancers are well suited for calibration and benchmarking of mutational analysis software, as there are frequently mutated genes with mutation hotspots (*KRAS* and *BRAF*), a tumor suppressor gene with well-known spectrum and prevalence of driver mutations (*TP53*), and a tumor suppressor gene inactivated by nonsense as well as frameshifting indel mutations (*APC*). A subset of CRCs has mismatch repair deficiency, manifesting in large numbers of indels in polymeric base contexts, and are likely to respond to checkpoint inhibitor treatment. Furthermore, diagnostic mutational analyses of solid tumors currently rely on enrichment panels of 50-500 genes sequenced to ∼1000-fold depth, and occasionally the whole human exome at ∼200-fold depth. Therefore, we used the following three sequencing protocols for benchmarking: (i) a custom designed Haloplex panel (Agilent) of 676 genes (PCR-based target enrichment) sequenced on Illumina HiSeq 2000 (Haloplex/HiSeq), (ii) the TruSight Oncology 500 gene panel (hybrid-capture target enrichment) sequenced on Illumina NextSeq 550 (TSO500/NextSeq) and (iii) the TWIST core exome panel (PCR-free target capture) sequenced on Illumina NovaSeq 6000 (TWIST/NovaSeq).

### Efficient and robust software for clinical mutation analyses in cancer

Combinations of different, and independently developed software for read trimming, alignment and mutation calling remains the state-of-the-art for mutational analysis in many research and clinical laboratories. Two commonly used pipelines are Cutadapt-BWA-MuTect2 (CBSM) and Cutadapt- BWA-Strelka2 (CBSS) (Figure 1A and B). Tight integration between the sequential steps of read trimming, alignment and somatic mutation analysis is required to enable parallel and rapid execution to enable the user to assume full responsibility for the output. We therefore developed VARify, a rigorous software pipeline, *de novo* with sole focus on optimal tumor-normal mutational analyses of cancers (Figure 1C). A derivative pipeline, ALTOmate, was developed for situations where no patient-matched reference sample is available (Figure 1D). Both VARify and ALTOmate accept for input the standard single-read or paired-end FASTQ files from the major sequencing providers (Illumina, MGI, IonTorrent or Pacific Biosciences) and output mutation calls in VCF format. Both pipelines consist of three functionalities, namely read trimming functionality (RTF), read alignment functionality (RAF), and variant calling functionality, integrated in a single executable.

### Read trimming functionality

If adapter sequences are provided, the RTF of VARify and ALTOmate will perform 3′ adapter detection and trimming in paired-end mode. The output of the RTF closely matched the output from Cutadapt v.4.0 with disagreement in on average 1/10,000 processed reads (Suppl. Table 1A). The difference stemmed from the alignment models used prior to trimming; RTF performed full-length Smith-Waterman alignment, whereas Cutadapt relied on semi-global alignment (19). When compared in 107 tumor and normal samples from the CRC set sequenced in pair-end mode, the RTF output was identical to Cutadapt in average 99.98% of the reads (Suppl. Table 1A). Both agreed on the presence of adapter sequence in 17.66% of the remaining reads, but the trimmed sequence differed (Suppl. Table 1A). The proportion of trimming decisions identical for both software was similar for read1 and read2 (99.98% in both), despite the difference in the length of the adapters to align (34 and 58 nucleotides for the first and second adapter respectively), suggesting that the alignment length is not important for the adapter detection and trimming decision. The RTF aims to identify and remove most of the adapter sequences while preserving most of the original sequencing data. Therefore, following adapter trimming, the RTF preserved slightly more (0.003% on average) of the original sequence compared to Cutadapt (Suppl. Table 1A). The VARify RTF was more than three times as fast as Cutadapt v.4.0 (Suppl. Table 1C). Since efficiency can be a reason to use faster read-trimming tools in clinical practice, we benchmarked to the ultra-fast trimming solution fastp v.0.20.0 (35). In the CRC dataset, fastp discarded ∼4.09% of the read pairs (∼8% of the data) which did not pass its internal quality threshold, so a direct comparison between fastp and either Cutadapt or RTF resulted in low concordance. If considering only the reads processed by fastp, trimming decisions by RTF and fastp were identical in ∼96.15% of the reads (Suppl. Table 1 B). In the remaining 3.85% of the reads, the adapter was detected but processed differently in ∼3.90% of the reads, and in another ∼90.19% of reads, the trimming decision involved <=5 nucleotides by either software Thus, the total concordance between fastp and RTF in the reads processed by fastp was >99.62% (Suppl. Table 1B). The RTF had two times faster execution speed than fastp (Suppl. Table 1C). Together, a read trimming functionality superseding the current state-of-the-art in speed and amount of sequence retained, all while implementing conservative choices in the alignment model, was developed.

### Read alignment functionality

Next, the RTF output of VARify or ALTomate is processed by the RAF while in operational memory to avoid inefficient local or network storage operations. To evaluate its performance, we compared mapping and alignment to that of the BWA-MEM aligner (Suppl. Table 2A). Comparing total mapability in the CRC Haloplex panel set, the VARify RAF mapped on average 98.78% of the reads compared to 99.32% for BWA-MEM (Suppl. Table 2A). Interestingly, 63.94% of the average 0.37% of total reads mapped by BWA-MEM but not RAF had PHRED score 0, interpreted as a 100% probability that the alignment is wrong. On average 65.85% of the reads, which were mapped, paired and assigned to different chromosomes by BWA-MEM were not mapped by RAF. Alignment as a proper pair or singletons was reported by RAF in 21.83% of the inter-chromosomal BWA-MEM read pairs. From all mapped reads, RAF outputs 0.35% singletons compared to 0.10% for BWA-MEM. Similarly, proper pairs (98.9% in BWA-MEM vs. 98.19% in RAF) were not forced over non-proper pairs (1.14% in BWA-MEM vs. 1.81% in RAF). The concordance between BWA-MEM and RAF in reported mapping coordinates was on average 97.24%. When the mapping quality (MAQ) score of the concordant reads was compared, BWA-MEM assigned higher MAQ than RAF, with >5 PHRED units. The mean MAQ reported by BWA-MEM was 59.66 and by RAF 54.28 (Suppl. Table 2A). When excluding reads with MAQ of 0, meaning reads which are wrongly placed with 100% probability, BWA-MEM alignment missed on average more than six targets compared to RAF in every sample evaluated (Suppl. Table 2A). In all, the RAF aligned 0.5% less reads than BWA-MEM in the CRC set, primarily by not placing reads with ambiguous mapping or rejecting inter-chromosomal pairs. When performing alignment comparison in the TSO500 and TWIST datasets, the differential in mapability between RAF and BWA-MEM increased, however most of the reads mapped by BWA-MEM, but not RAF were with MAQ=0 (93% and 95% respectively). When excluding the zero quality reads, BWA-MEM missed on average more targets in comparison to RAF, 21 in the TSO500 and 1496 in the TWIST datasets (Suppl. Table 2B and 2C). Together, the RAF is more conservative than BWA-MEM as it avoids assigning ambiguous pairs and reads.

### Variant calling functionality

For benchmarking of the variant calling functionality of VARify, we compared pipeline performance on data from 676 genes sequenced in 82 MSS and 21 MSI CRC cases using Haloplex gene panel enrichment and Illumina HiSeq 2000 sequencing. When analyzing the somatic mutations reported for each sample by the three different pipelines, consecutive filters were applied to facilitate the interpretation. The CBSS, CBSM and VARify reported on average 4,318, 779 and 331 total mutations, of which 4,252, 753 and 321 on-target mutations, and 602, 190 and 208 mutations on average, respectively, passed all pipeline filters (PASS) (Suppl. Table 3). The calls of the three pipelines over CRC driver genes sequenced by the Haloplex panel were collated (Figure 2A and Suppl. Figure 1A). Thus, there is an overall consensus between the report of the three pipelines, with a tendency towards overcalling by CBSS and good agreement between CBSM and VARify.

**Figure 2.**
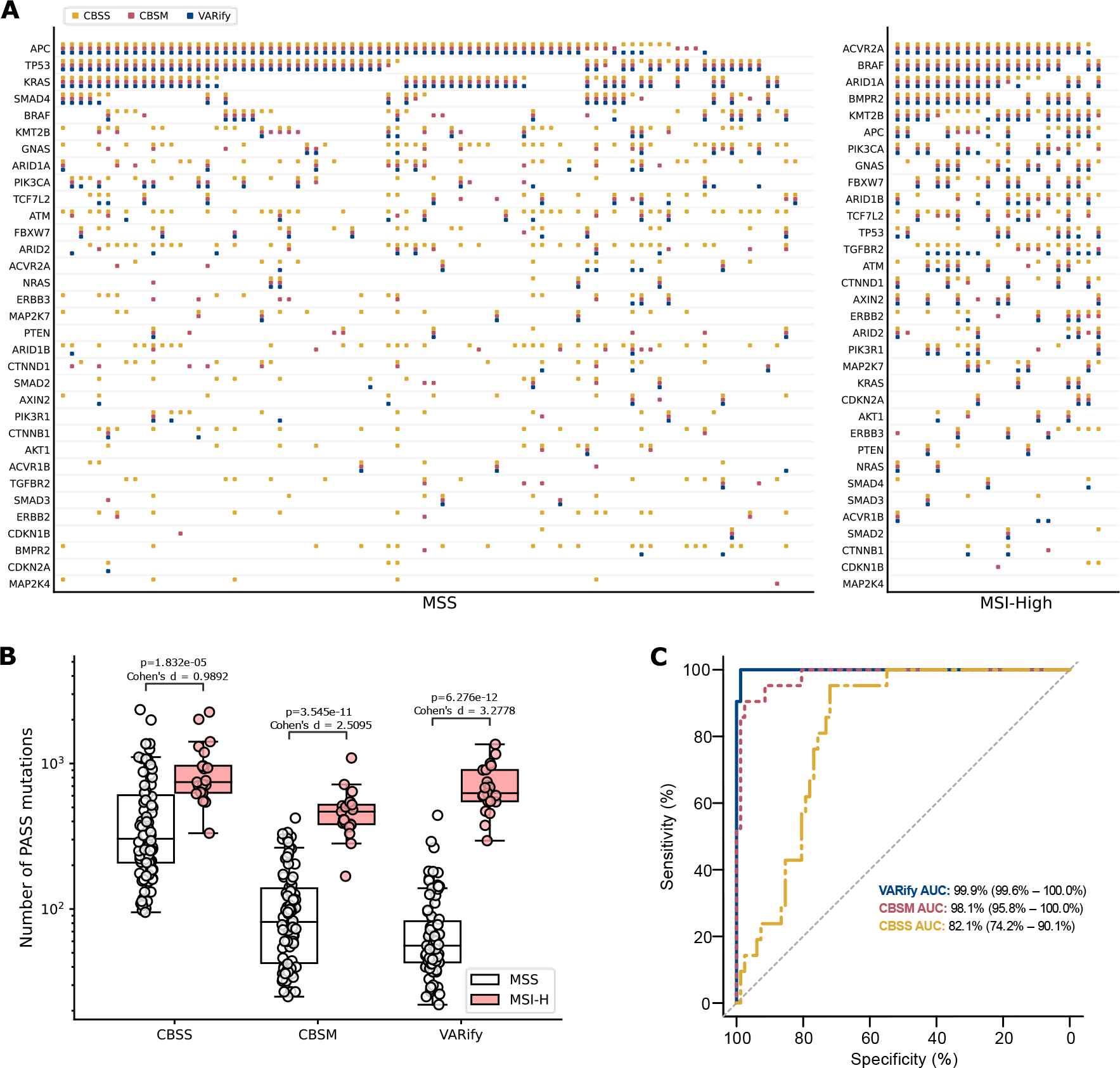
Accurate mutation calling and superior distinction of microsatellite instability in colorectal cancer by VARify. A total of 676 genes in 82 microsatellite-stable (MSS) and 21 microsatellite instability-high (MSI-High) colorectal cancer cases were enriched using a custom Haloplex panel and sequenced on Illumina HiSeq 2000. (**A**). The calling consensus over the top 33 driver genes for CRC, according to the mutation reports of CBSS (yellow), CBSM (magenta) and VARify (blue). (**B**). The microsatellite instability (MSI) status was determined using MSI Analysis System, v1.2 (Promega). The tumors were plotted in two bins according to their MSI- status; microsatellite stable (MSS) (open circles), and microsatellite instable MSI-High (salmon circles). The number of PASS mutations by each pipeline is plotted on a logarithmic scale. The targeted sequencing space was 5,240,231 bases. The p-values of Mann-Whitney U test with Bonferroni correction for comparison of the MSS and MSI-High sets is shown by pipeline, CBSS, CBSM and VARify. (**C**). The ROC-curve analysis of classification into MSS and MSI-High based on the number of PASS mutations, for CBSS (yellow), CBSM (magenta) and VARify (blue).

### Classification of CRC as MSI or MSS and determination of tumor mutation burden

The total number of mutations reported by CBSM and VARify, but not by CBSS, correlated with MSI status of the sample (CBSS p = 2.568e-01 with Cohen’s d = -0.4057, CBSM p = 5.881e-10 with Cohen’s d = 2.2643, VARify p = 5.930e-12 with Cohen’s d = 3.6573, Mann-Whitney U test) (Suppl. Figure 1B). If considering only mutations passing all filters (PASS), there was apparent statistical significance for all pipelines, but CBSS suffered from effect size, as suggested by Cohen’s d (Figure 2B). Further, these correlations improved if only the number of PASS InDels were compared (CBSS p = 4.435e-12 with Cohen’s d = 3.9200, CBSM p = 5.018e-12 with Cohen’s d = 3.9728, VARify p = 5.449e-12 with Cohen’s d = 3.5759) (Suppl. Figure 1C). The ROC analysis of MSI status classification by each pipeline revealed that CBSS and CBSM gradually improved when total mutations, PASS mutations or only PASS InDels were considered. VARify had the highest sensitivity and specificity when the total and PASS mutations were used for classification (Suppl. Figure 1D Figure 2C). When only the PASS InDels were considered, all three pipelines accurately classified the samples (Suppl. Figure 1E). Thus, VARify provided superior classification of MSI and MSS tumors as compared to other commonly used pipelines.

### Mutational analysis of cancer genes

In a manually-curated set of previously reported somatic mutations (27), the overall recall was 96%, 94% and 91% for VARify, CBSS and CBSM pipelines. The mutations observed in *KRAS* were mainly located in known hotspots. For all samples having sufficient read depth for analysis (coverage ≥30 reads), all KRAS mutations in codon 12, 13 and 61 observed by pyrosequencing (36) were detected by all pipelines. Two transforming hotspot mutations at p.A146T (37), an oncogenic p.T74P mutation (37, 38), an oncogenic p.K117N mutation (37, 39), a potentially pathogenic p.D119H mutation (40, 41) and several novel mutations (p.E91D, p.A11D and p.I93N) were also detected by VARify. In *BRAF*, all V600E mutations detected *a priori* by pyrosequencing and covered with ≥30 reads were reported by VARify (27). Driver somatic mutations in *TP53* have been characterized and annotated (42) and 70% of CRC cell lines had non-synonymous or truncating mutations (43). Here, 63% of the CRC tumors had at least one *TP53* mutation in the protein coding sequence, which is in agreement with prior knowledge (44), and 97% of non-synonymous, nonsense and indel mutations reported here had previously been reported in the UMD *TP53* database (45). Of the latter, 73% were missense, 15% nonsense, and 11% frameshifting indels, in agreement with previous reports (44, 46). In 82% of the MSS cases, *APC* had inactivation mutations. Of these, 55% (45/82) had one mutation and 45% (37/82) had two different mutations, with 80% of all mutations located in the mutation cluster region (MCR) and 98% of total observed somatic alterations being truncating (41 small indels and 55 stop-gain SNVs). The distribution and prevalence of somatic *APC* mutations was in agreement with previous reports that the tumor suppressor *APC* is inactivated by somatic mutations in the vast majority of MSS CRC cases with a proportion of small indels to stop-gain SNVs near 1:1 and the majority of mutations observed in the mutation cluster region (47, 48). Of the nonsense mutations, 92% were previously reported in the UMD *APC* database (49) or COSMIC (50).

Together, the results from analyses of known cancer genes demonstrate the ability of VARify to accurately detect driver somatic SNVs as well as indels in deep sequenced human solid tumors.

### Consistency of mutation reports across different sequencing and enrichment technologies

One strategy to validate true somatic mutations and exclude sequencing artifacts is to subject the same sample to conceptually different enrichment and sequencing technologies. To probe the calling consistency of the three pipelines over three different library preparations and sequencing instruments, we used materials from the same DNA extraction performed on 4 pairs of tumor and patient-matched normal samples (MSS-1, MSS-2, MSI-Low and MSI-High). Consistent calling with sufficient sensitivity over all three panels required that the evidence for a prospective call by VARify exceeded 2 PBR units. Overall, the three pipelines and sequencing platforms agreed on very few somatic events in exons covered by all three libraries (Suppl. Table 4). In the intersection of the sequencing targets for each library, all pipelines reported the same 4 mutations in MSS-1, 5 mutations in MSS-2, 17 mutations in MSI-Low and 31 mutations in MSI-High (Table 1 and Suppl. Table 4). Although VARify is agnostic to target definitions, it reports the fewest total mutations (Suppl. Table 3) and very few mutations outside of the consensus with 3, 7, 12 and 5 mutations for MSS-1, MSS-2, MSI-Low and MSI-High respectively (Figure 3A). In comparison, CBSM had tens of additional calls (12, 53, 22 and 27), while CBSS had hundreds (343, 518, 289 and 293) (Figure 3A). The core consensus set of mutations per sample (Figure 3A) represents the molecular profile of driver events in MSS and MSI tumors, and the numbers of mutations correlate with the subtype of CRC (Table 1). The number of reported mutations by VARify correlated with the MSI status of the sample, and VARify reported the least putative calls compared to CBSM and CBSS. In a side-by-side comparison, VARify was faster than CBSM or CBSS pipelines in the Haloplex set and faster than CBSM in TSO500 and Twist exome analysis (Figure 3B). The speed enhancement primarily stems from tight integration of the different functionalities, elimination of intermediate steps, and multiprocessing. Together, VARify produced consistent results with less outside the consensus of the three pipelines in the same target across different sequencing technologies.

**Figure 3.**
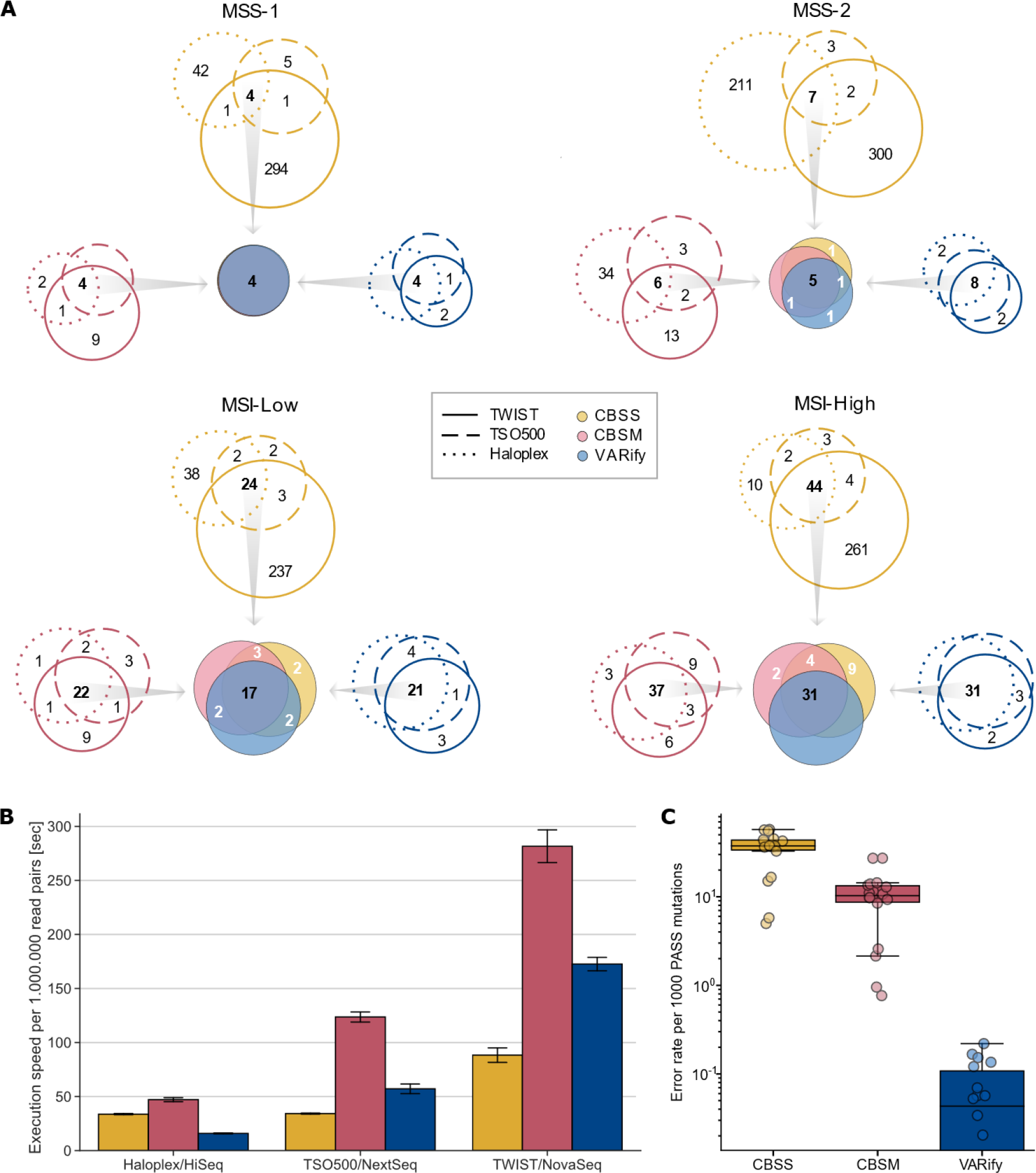
The VARify pipeline provides rapid, specific and consistent somatic mutation detection across three different sequencing platforms and panel enrichment technologies. (**A**) The consensus of each of the three pipelines evaluated (CBSS, CBSM and VARify) in the intersection of the target space for Haloplex/HiSeq (⸱⸱⸱), TSO500/NextSeq (---) and TWIST/NovaSeq (⸻) in four colorectal cancer DNA samples from tumors with different microsatellite instability phenotype (MSS-1, MSS-2, MSI-Low and MSI-High). The consensus of each pipeline over the three sequencing platforms is compared to the consensus of other pipelines in each central Venn (filled). CBSS (yellow), CBSM (magenta) and VARify (blue). (**B**) The average processing time for one million read pairs from 4 samples sequenced with Haloplex/HiSeq, TSO500/NextSeq and TWIST/NovaSeq and analyzed with CBSS (yellow), CBSM (magenta) and VARify (blue). Mean and SD. (**C**) The error rate determined as the number of erroneous mutations per 1000 PASS positions reported by CBSS (yellow), CBSM (magenta) and VARify (blue) pipelines in the 18 WES samples of the SECQC2 dataset (logarithmic scale).

**Table 1.**
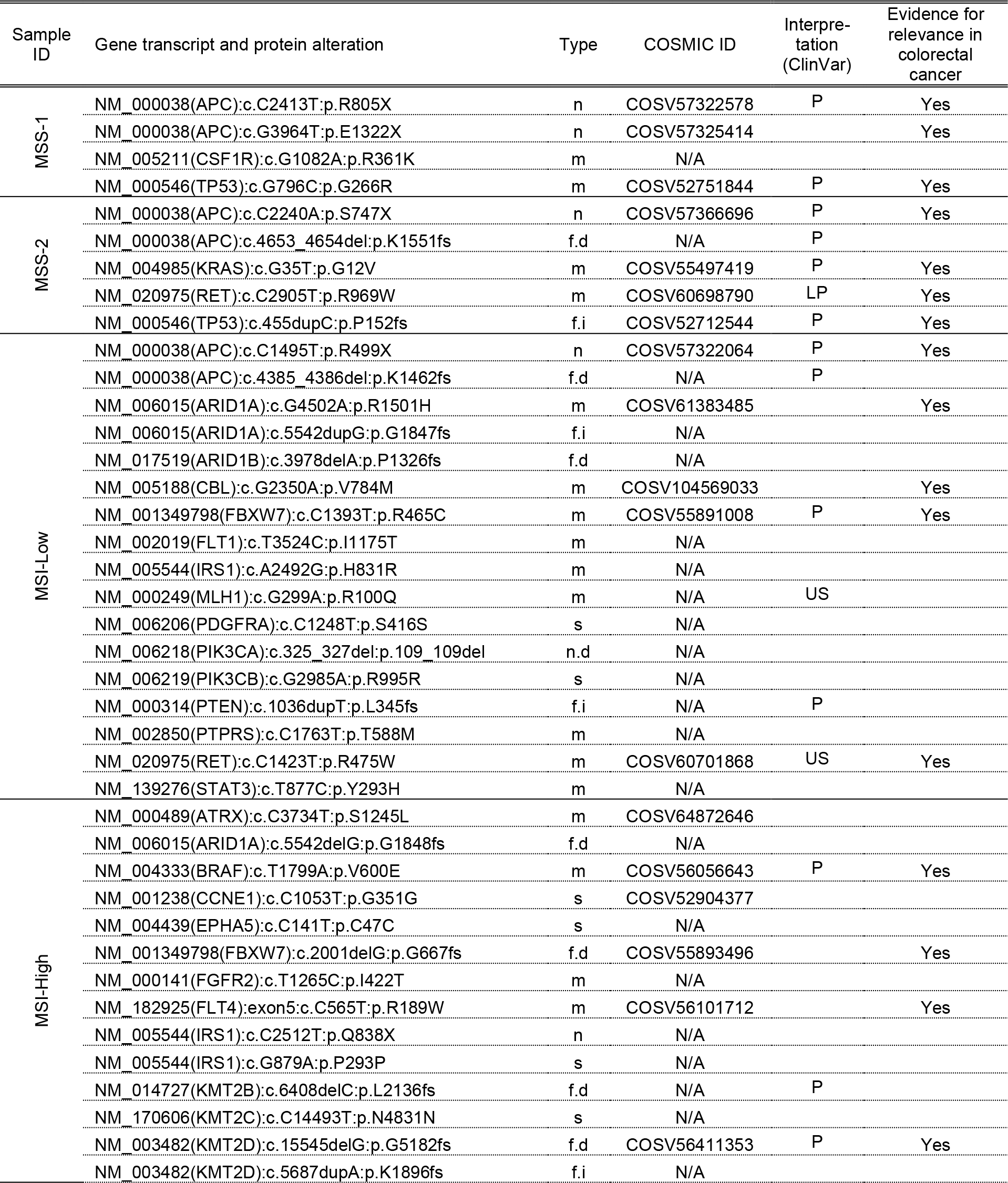
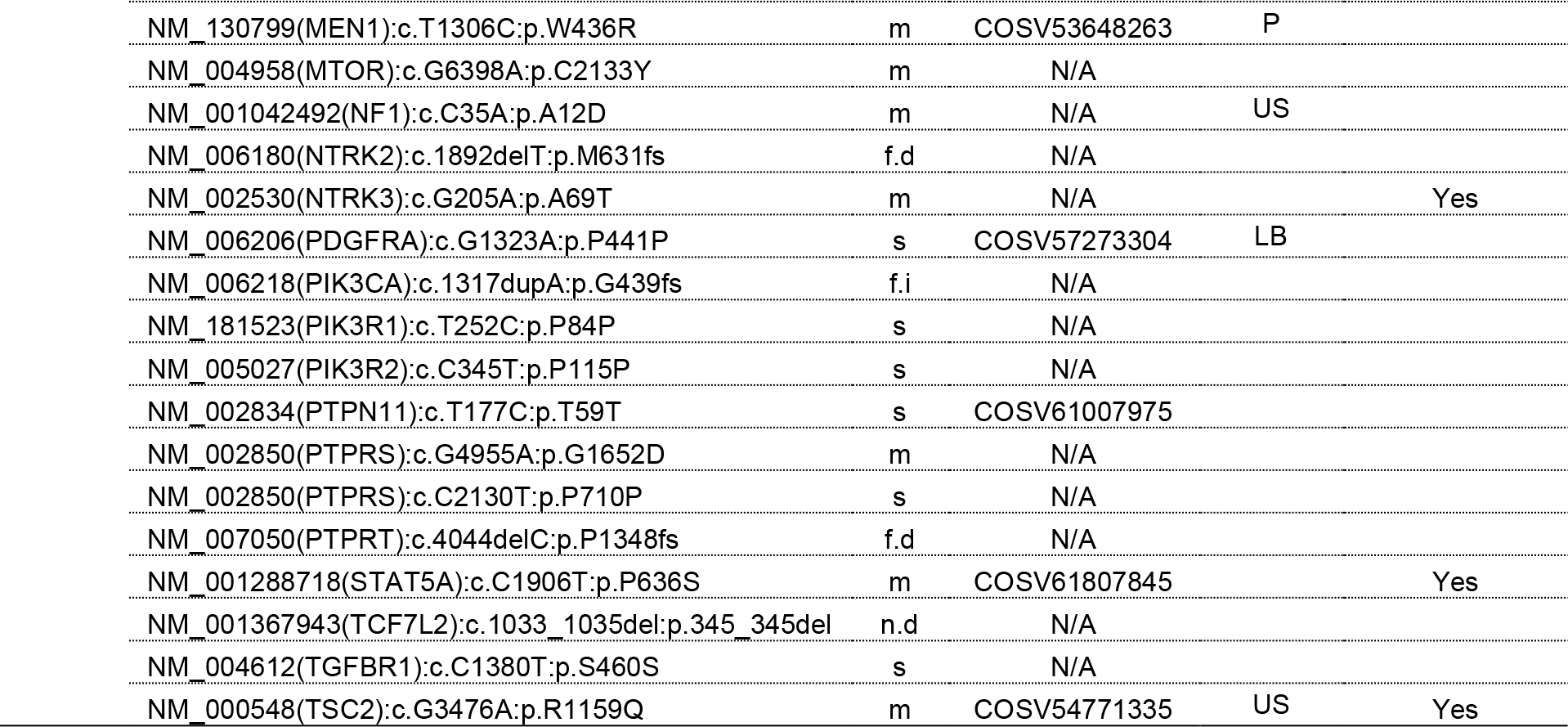
Mutation consensus reported by CBSM, CBSS and VARify pipelines in four samples sequenced by Haloplex/HiSeq, TSO500/NextSeq and TWIST/NovaSeq. For each sample pair, the alteration in the gene transcript and protein sequence is shown, together with the genome coordinates of the mutation (GRCh37 as reference), the reference and the alternative allele, type of mutation (n, nonsense; m, missense; f.d, frameshift deletion; f.i, frameshift insertion; n.d, non-frameshift deletion; s, synonymous), COSMIC ID (if available) and the interpretation in ClinVar database (P, Pathogenic; LP, Likely Pathogenic; LB, Likely Benign; US, Unknown Significance).

### Quantification of false positive and true positive mutation calls

The appearance of mutations in normal-normal calling can be explained by somatic mosaicism or artifacts in re-sequencing the same sample (51). The idea of using technical replicates of a NGS sequenced normal sample for evaluation of somatic mutation performance is in pre-print (52). The SEQC2 dataset is a manually curated reference for identification of false positive and true positive calls (30), that we used here. To eliminate biological sources of variants in the scoring of false positive mutations, we first limited the analysis to pairs of identical data. Thereby, the calling test directly scores false positive somatic mutations, with a baseline at zero. Calling a SEQC2 aNormal-Normal data pair, VARify reported no somatic mutations while the CBSM pipeline reported on average 120,411 putative events of which average 1,041 were PASS somatic mutations and the CBSS pipeline reported 200,219 putative events scored as possible somatic mutations with the LowEVS filter (Suppl. Table 5A). The CBSM yielded different output when analyzing aNormal vs Normal or Normal vs aNormal, whereas the CBSS and VARify output was independent of the direction of the pair. Many of these erroneous calls were present in the tumor-normal calling of the same pair, average 9,879 for CBSS and 13,332 for CBSM (up to 8 PASS) (Suppl. Table 5B). Finally, we performed Normal-Normal calling without any alteration with samples that come from different sequencing experiments, resulting on average in 57,638, 19,310 and 600 PASS mutations for CBSS, CBSM and VARify respectively (Suppl. Table 5C). The CBSS, CBSM and VARify pipelines were compared with respect to the false positive mutations in manually curated SEQC2 dataset (Suppl. Table 6). False positive calls were defined as all reported mutations with genome coordinates falling in the list of known negative positions evaluated by SEQC2. On average CBSS, CBSM and VARify reported 2,282,443, 970,491 and 129,598 total calls per sample, from which respectively 48,037, 44,569 and 15 were in the known negative list. If considering only PASS calls, CBSS, CBSM and VARify reported on average 508,125, 186,636 and 60,112 calls, with 19,151, 1,760 and 4 in the known negative list. In addition to the 284 putative calls for the SEQC2 dataset, 39 on-purpose selected true-negatives were also probed by ddPCR. The mutation report of CBSS, CBSM and VARify did not include any true negatives. All pipelines reported 220 true positives, however there was a difference between the pipelines in reporting of the 64 false positive calls. Respectively, CBSS and CBSM reported in total 57 and 3 of the false positive mutations (none were PASS), while VARify did not report any false positive event either in the raw or the PASS output (Figure 3C). Taken together, VARify produces less false positive mutation calls than the state-of-art.

### Recall of the consensus mutations in IonTorrent data

The CBSS and CBSM pipelines are not optimized to handle ion semiconductor data and benchmarking to them was therefore not possible. However, it was possible to employ ALTOmate, the tumor-only derivative pipeline of VARify, when only the tumor sample is sequenced. Although the target space for Illumina and IonTorrent technology differed, VARify and ALTOmate re-called all mutations in the cross-section in the IonTorrent data from the four pairs of tumor and patient-matched normal samples (Suppl. Table 7). Thus, ALTOmate and VARify can be applied also to analysis of IonTorrent data.

## Discussion

In the diagnostic setting, high precision is equally important as high sensitivity. However, most software for mutational analyses was originally developed for scientific use in constitutional genomics where sensitivity may be more of a priority than specificity. This leaves the diagnostic specialist, e.g. the molecular pathologist, with uncertainty as to the reliability of mutation detection and thereby tends to limit the interpretation of sequence data outside of established hotspot or recurring mutations in the case of cancer specimens. The demand on sequencing technologies for more uniform and higher average sequencing depth is likely to provide better calling breadth and sensitivity, but also demands high precision for the mutation calling (53, 54). A clinical report with fewer false positive mutations requires a shorter time for interpretation and validation. Therefore, the goal of the current clinical protocols is to enhance precision, while keeping high sensitivity. VARify provides such a balance between high precision and high sensitivity, reporting the disease-relevant mutations with the lowest background of false-positive mutations.

Mutation interpretation and therapy selection should rely on a robust and consistent mutation report across different sequencing panels and technologies, without loss of sensitivity. Here, we compared three separate gene panels and sequencing protocols and demonstrated higher consistency of the VARify mutation report compared to CBSS and CBSM. While all three pipelines uncovered the key CRC driver mutations, the background in CBSS and CBSM was two orders of magnitude higher than VARify. This translates to reduced efforts in manual removal of false positive mutations using VARify. The false positives mutations generated by CBSS and CBSM were evident in the analysis of identical sequencing datasets, whereas VARify did not produce such calls. Further, CBSM showed an unexpected stochastic behavior, with differences in the mutation reports when the direction of the identical samples was mirrored, which is undesired in a clinical setting where reproducibility is crucial. To evaluate the impact of false positives in the analysis of identical sequencing data by CBSS and CBSM, the calls were traced in the output of a tumor-normal analysis. While in the mutation report of CBSS all such events were filtered out, several false positive mutations reported by CBSM passed all filters. This fact makes it imperative to run identical dataset analysis by CBSS and CBSM side by side to the real tumor-normal analysis to remove such artifacts, while the analysis of VARify does not report this class of false positive mutations. When two independent sequencing experiments of the same normal DNA sample were compared there are calls to be expected due to somatic mosaicism or technical artifacts (51). However, in favor of VARify, the number of reported events passing all filters differed in several orders of magnitude between the pipelines. Deviations in each analyzed sample pair from the average number of passed mutations may be attributed to the quality of a sequenced samples, however, the general tendency in the number of reported events was consistent for each pipeline. A substantial part of these calls is due to somatic mosaicism, sequencing artifacts or limitations in the analysis, but the number of such calls were 11.3%, 10.3% and 1% of the PASS mutations for CBSM, CBSS and VARify respectively, indicating that the vast majority are false positive mutation calls. Combined with the larger mutation outputs of CBSS and CBSM in comparison to VARify, the number of putative events may require a large validation and tedious manual curation effort.

Immunotherapy treatment decisions incorporate information from complex biomarkers, such as TMB and MSI status. However, correct interpretation requires an accurate mutation report, where the false positives currently constitute a huge obstacle. Here, VARify stratified tumors by MSI status already in the raw unfiltered mutation report, and still outperformed the alternative pipelines using filtered output. If only the small base insertions and deletions are considered, all pipelines agree, which poses a question if the substitutions are relevant for such analyses. The correlation between the total number of mutations and the MSI status suggests that substitutions are not critical for the analysis by VARify, although somewhat challenging for CBSS and CBSM at this relatively small sequence target of ∼5,2 Mb. Together, it is evident that the omission of substitutions improves the correlation for all pipelines in this target space.

The VARify pipeline is based on comparison of T/N samples, which are often not available in a clinical setting, or if available, not sequenced due to higher cost and perceived requirement of double the analysis per patient. The higher cost of performing two analyses can potentially be justified by the reduced cost of treating patients with targeted therapies because of false positive mutations. Another current challenge for clinical laboratories is to analyze and compare genetic data based on several different sequencing platforms, often including legacy technologies. Current diagnostic protocols rely on ion semiconductor sequencing or sequencing by synthesis, which in turn rely on analysis software that supports the one, but not the other technology. The CBSS and CSSM pipelines are tailored towards paired-end sequencing by synthesis, while the IonReporter pipeline only functions with the native single-read mode of the ion semiconductor sequencing. In contrast, both VARify and ALTOmate successfully processed data from both sequencing technologies. The long-read sequencing technologies of Pacific Biosciences or Oxford Nanopore have great potential to answer complex clinical questions beyond the somatic SNV calling and may enter routine practice in the coming years. VARify have been designed with long-read data in mind, and the underlying algorithms were already exploited for PacBio analysis in a research setting (55). Thus, VARify and ALTOmate can utilize the input from various sequencing vendors to produce accurate mutation reports in a format suitable for clinical interpretation, streamlining of clinical sequencing pipelines and facilitating inter- or intra-lab comparisons for a robust cancer precision medicine.

## Data Availability

All data produced in the present study are available upon reasonable request to the authors

## Acknowledgements

This work was funded by grants from the Swedish Agency for Innovation Systems (Vinnova) and EU FP7 (MERIT). The authors wish to thank Dr. Chatarina Larsson for critically reviewing the manuscript. The use of PBR in this and other applications is governed by patents WO2016043659A1 and US10325183B2. IS, TA, IC, VL and TS are co-founders of Oncodia AB which holds all rights to VARify and ALTOmate.

---

**Supplementary Figure 1.**
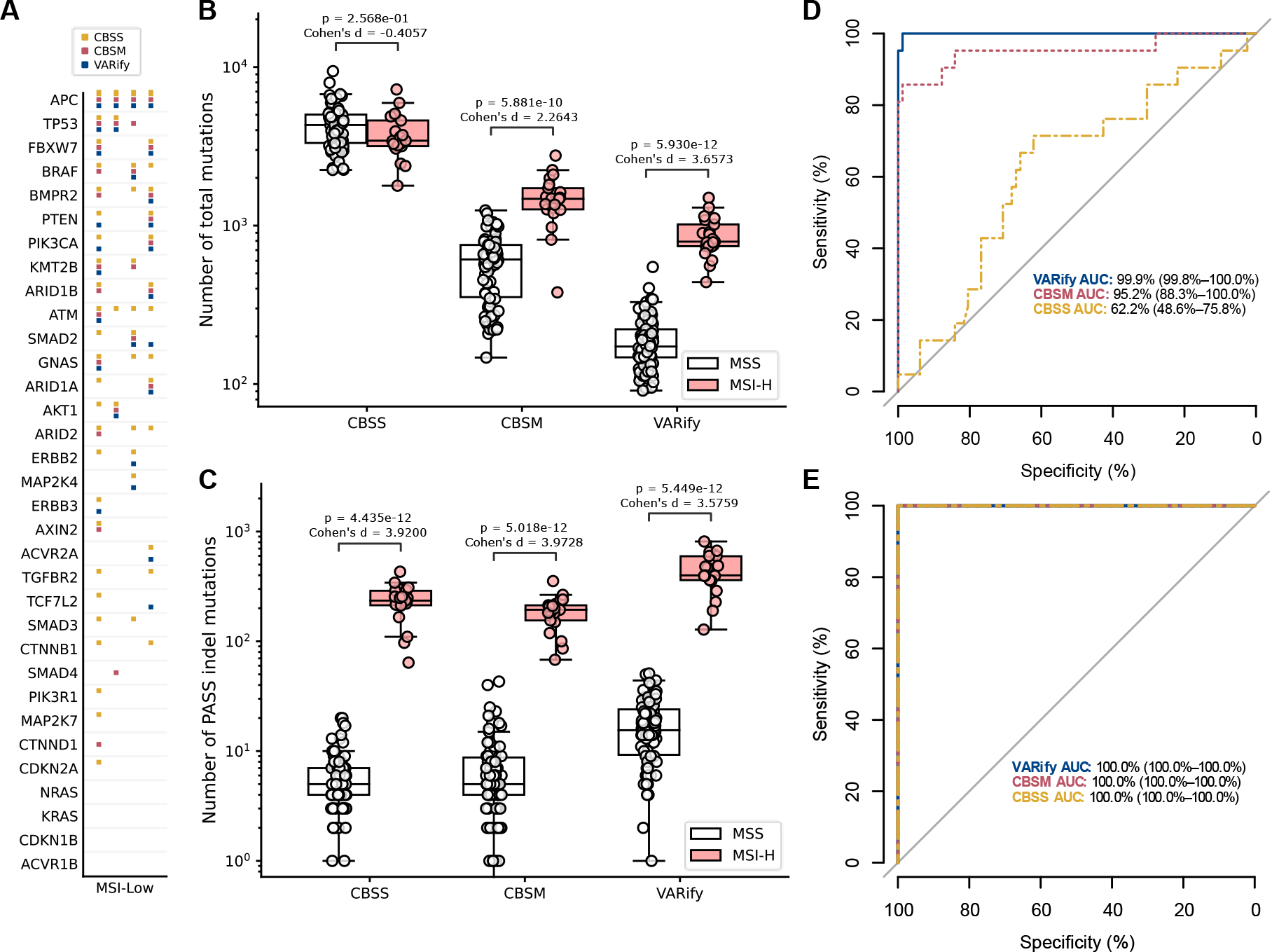
The total mutation report of VARify is sufficient to accurately classify CRC samples by their MSI status. DNA of 107 CRC cases was enriched for a set of 676 genes using a custom Haloplex panel and sequenced on Illumina HiSeq 2000. The microsatellite instability (MSI) status was determined using MSI Analysis System, v1.2 (Promega). (A) The calling consensus over the top 33 driver genes for CRC in four microsatellite instability-low (MSI-Low) samples, according to the mutation reports of CBSS (yellow), CBSM (magenta) and VARify (blue). The total number of reported mutations (B) and the number of InDels passing all filters (C) by each pipeline is plotted in a logarithmic scale. The p-values of Mann-Whitney U test with Bonferroni correction for comparison of the MSS and MSI-High sets is shown by pipeline, CBSS, CBSM and VARify. The ROC-curve analysis of classification into MSS and MSI-H based on the total number of mutations (D) and the number of InDels passing all filters (E), for CBSS (yellow), CBSM (magenta) and VARify (blue).

## Notes

### Author Declarations

The mutational analyses of patient samples were approved by the Ethical Review Board in Uppsala (EPN Uppsala, C116/2007).

